# Electrical Stimulation and Time to Radiographic Healing of Acute Fractures: A Systematic Review and Meta-Analysis

**DOI:** 10.1101/2021.05.09.21256916

**Authors:** Peter J. Nicksic, Kevin Rymut, Aaron Dingle, Nishant Verma, Christopher Doro, Andrew Shoffstall, Kip Ludwig, Samuel O. Poore

## Abstract

**Objective:** To determine if electrical stimulation (ES) reduces days to radiographic union of acute fractures

**Data Sources:** MEDLINE database search using the terms combinations of “electric stimulation AND bone healing”, “electric stimulation AND fracture,” “electric stimulation AND fracture healing,” full articles, English language, without publication date restriction

**Study Selection:** Inclusion criteria were (1) randomized-controlled trials concerning electrical stimulation for the purpose of healing acute fractures with (2) outcomes on radiographic union at regular time intervals. Exclusion criteria were (1) studies involving skeletally immature patients or (2) ES for non-unions, spinal fusions, and osteotomies.

**Data Extraction:** Study quality was assessed with the Cochrane Collaboration tool for risk of bias assessment by 2 independent reviewers. Heterogeneity between studies was assessed with the χ^2^ and I^2^ tests.

**Data Synthesis:** The mean days to radiographic union was calculated as a continuous variable with standard deviations. The meta-analysis was performed to compare the ES and non-ES groups across studies using Metafor ® software (Bell Labs, Murray Hill, NJ, United States).

**Conclusion:** Electrical stimulation does not reduce time to radiographic union in acute fractures. However, an improvement in the healing time was noted in the semi-invasive method of ES in which the current was delivered directly within the fracture site. This finding provides evidence that innovative methods of ES delivery may demonstrate the promising results found in smaller animal studies.

**Level of Evidence:** Level I

## Introduction

With approximately 18.3 million cases in the United States^1^ per year, orthopedic fractures are extremely common and represent a large public health burden in terms of cost and socioeconomic impact secondary to work disruption and disability. The total economic impact of orthopedic fractures has been estimated at $265.4 billion dollars per year.^1^ Further, delays in healing incur significant morbidity to the patient in the form of pain and functional limitations.^2^ The time required for fractures to unite is important because of the resources used to care for patients until they are independent again as well as the amount of lost wages from being off work. Understanding and expediting the process of fracture healing would greatly improve patient care.

One method that has been described to augment bone healing is electrical stimulation (ES). Since Fukada and Yasuda first described the piezoelectric property of bone – that bone generates endogenous electrical fields by ionically-driven currents in response to mechanical stress – and its relationship to bone formation,^3^ there has been an abundance of evidence supporting the use of exogenous ES to augment bone formation in both the *in vitro* and *in vivo* animal model.^4-6^ However, the clinical data in support of ES remain mixed. The lack of consensus in the literature may be due to differences in definition of radiographic union, varied fracture location and type, poor patient compliance with device utilization, or issues with delivery of ES to the fracture site.

Methods utilized to deliver ES to fractures include invasive (or semi-invasive) current, capacitive coupling, and inductive coupling. Invasive current is a method of delivery in which there is a cathode that is placed through the skin in or near the fracture site. Non-invasive ES delivers weaker, time-varied electromagnetic fields reaching the fracture site with capacitive or inductive coupling. Capacitive coupling devices use electrical fields transmitted transcutaneously though pads placed on the patient to generate current at the fracture site. Non-invasive inductive coupling ES includes pulsed electromagnetic field (PEMF) and combined magnetic field (CMF). PEMF is commonly used and described in the literature and typically involves coils placed on the patient to transcutaneously generate electromagnetic treatment fields.

There have been two recent meta-analyses studying the efficacy of ES for the purposes of bone healing in non-unions and spinal fusion,^7,8^ which demonstrate that the relative risk of non-union at one year is reduced by 35% in the ES group.^8^ However, while these meta-analyses focused on studies comparing rate of non-union – a fracture that fails to heal within the period of time that is considered ‘normal’ for the particular fracture, none to date have compiled data exclusively on the effect of ES on time to healing of acute fractures. With the aim of answering the question if ES decreases the days to radiographic union of acute fractures, we performed a systematic review and meta-analysis of randomized-controlled trials with the outcome of interest being the days to radiographic union.

## Methods and Materials

Utilizing the standard PRISMA protocol,^9^ a MEDLINE database search was conducted on September 25, 2020, using combinations of search terms of “electric stimulation AND bone healing”, “electric stimulation AND fracture,” “electric stimulation AND fracture healing,” “direct current AND bone healing”, “inductive coupling AND bone healing,” and “pulsed electromagnetic frequency AND bone healing.” The complete search strategy is included in **Appendix 1**. Additionally, all bibliographies of systematic reviews and meta-analyses on the topic were searched for articles that may have been missed in the MEDLINE search. There was no date of publication restrictions. Inclusion criteria were randomized-controlled trials (RCTs) concerning ES for the purpose of healing acute fractures of the appendicular skeleton with outcomes on radiographic union at regular time intervals. Studies involving skeletally immature patients or ES for non-unions, spinal fusions, and osteotomies were excluded. Two reviewers (P.J.N. and K.R.) agreed upon inclusion of studies for meta-analysis after full-text review. Data for article title, authors, year of publication, study type, number of patients studied, fracture type, ES type, fixation method, ES protocol, ES device specifications, radiographic union definition, radiographic interval, length of follow up for each group, number of patients lost to follow up for each group, non-unions in each group, and days to radiographic union was collected by each reviewer, and disagreements were handled through discussion or inclusion of a third reviewer (S.O.P) as a tiebreaker. For each study, the reviewers (P.J.N and K.R.) independently assessed the study quality per the Cochrane Collaboration tool for risk of bias assessment.^10^

The outcome of interest was time to radiographic union. This was recorded as a continuous variable in days, with means and standard deviations calculated. Non-unions requiring secondary surgery were included in the analysis as a union at the longest measured time-point in each study. For example, if a scaphoid fracture went on to non-union and required open reduction, internal fixation (ORIF) with bone grafting at 3 months, this fracture would be counted as reaching radiographic union at 1 year, which is the longest time to union recorded in the study. This was done to capture the clinically relevant non-unions in the analysis. Heterogeneity between studies was tested using both the χ^2^ test (significance defined as p < 0.05) and the I^2^ tests (substantial heterogeneity defined as values >50 %).^11^ The studies were also analyzed in subgroups for upper and lower extremity location of fracture and operative – ORIF, external fixator placement, or percutaneous pinning – and non-operative – closed reduction and casting – methods of fracture fixation. The meta-analysis was run on Metafor ® by the Department of Surgery biostatistician group.^12^

## Results

Our query yielded 1652 references (dates ranged: 1964-2020). After title and abstract screen, 27 studies underwent full-text analysis. Five clinical trials and 301 patients were included in the meta-analysis (**Figure 1**). Of these 5 trials, two were studies pertaining to fractures of the lower extremity,^13,14^ and three were for fractures of the upper extremity.^15-17^ There were two studies concerning non-operative fractures,^15,16^ and three studies for operative fractures.^13,14,17^ There were four studies that used PEMF as their mode of stimulation,^13-16^ and one study used continuous current.^17^ Martinez-Rondanelli *et al*.^14^ and Faldini *et al*.^13^ utilized a removable transcutaneous PEMF device – a device that the patient applied to the affected extremity daily. Faldini *et al*.^13^ used stimulators with 75Hz, 1.3μs impulse duration, and 2mT peak magnetic field, where as Martinez-Rondanelli *et al*.^14^ did not report specific values. Hannamen *et al*. 2012^16^ and Hanneman *et al*. 2014^15^ used cast-implanted transcutaneous PEMF devices – a device that was not removable and was implanted in the thumb spica cast. For both studies, the stimulators were programmed for 50mV pulse amplitude, 5μs pulse width, 5ms burst width, 62ms refractory period, and 15Hz repetition rate.^15,16^ Finally, Itoh *et al*.^17^ administered continuous current through the pin of an external fixator directly into the fracture site of intra-articular distal radius fractures (2Hz, 30μA, 0-60kΩ). As noted above, this study was the only one we reviewed which utilized a semi-invasive means for stimulation – that is, ES delivered directly within the fracture site. The studies analyzed, including the specific methodology of ES, are summarized in **Table 1**.

**Figure 1:**
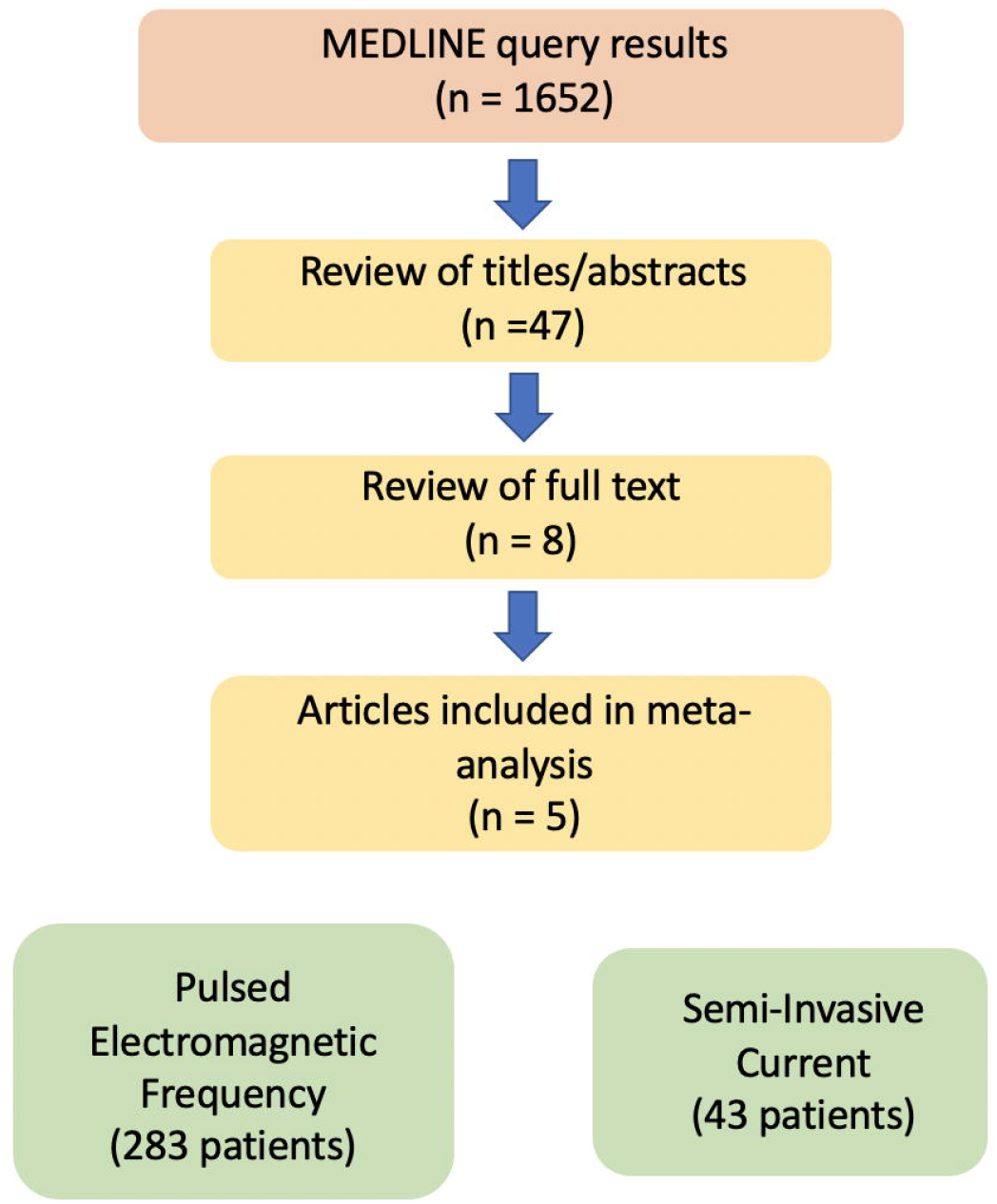
Flowchart of records included in the meta-analysis. A MEDLINE search was conducted per PRISMA protocol for relevant key terms to include randomized, sham-controlled trials for ES of acute fractures with time to radiographic union as a measured outcome. After applying all review criteria, our analysis included n=5 papers, with a total 301 patients. Four studies used non-invasive methods of stimulation, and 1 study used an invasive means of delivering current directly into the fracture site through the pin of an external fixator.

**Table 1:** The 5 included studies with details regarding methodologies, including fracture location and operative invention, type of ES, ES regimen, treatment arms in the study, and definition of radiographic union.

| Author | Title | Journal | Year | Study Type | Fracture Location | Operative Fracture | Groups | ES type | ES regimen | Union definition |
| --- | --- | --- | --- | --- | --- | --- | --- | --- | --- | --- |
| aldini et al. | Electromagnetic bone growth stimulation in patients with femoral neck fractures treated with screws: prospective randomized double-blind study | Curr Orthop Pract | 2010 | RCT | Femoral Neck | Yes | ORIF w/ sham vs. ORIF w/ ES | PEMF | 8hr/day for 13 weeks | 70% trabeculae bridging |
| anneman et al. | The clinical and radiological outcome of pulsed electromagnetic field treatment for acute scaphoid fractures | J Bone and Joint Surg | 2012 | RCT | Scaphoid | No | Cast w/ sham vs. cast w/ ES | PEMF | 24hr/day for 6-12 weeks | Signs of trabeculae bridging on XR views |
| anneman et al. | CT scan-evaluated outcome of pulsed electromagnetic fields in the treatment of acute scaphoid fractures | Bone and Joint J | 2014 | RCT | Scaphoid | No | Cast w/ sham vs. cast w/ ES | PEMF | 24hr/day for 6 weeks | 75-100% trabeculae bridging |
| Martinez-ondanelli et al. | Electromagnetic stimulation as coadjuvant in the healing of diaphyseal femoral fractures: a randomized controlled trial | Colombia Medica | 2014 | RCT | Femoral diaphysis | Yes | IMN w/ sham vs. IMN w/ ES | PEMF | 1hr/day for 8 weeks | Partial or complete union per t radiologi |
| Itoh et al. | Treatment of distal radius fractures with a wrist-bridging external fixation: the value of alternating electric current stimulation | J Hand Surg | 2008 | RCT | Distal Radius | Yes | Spanning fixator vs. spanning fixator w/ ES | CC | Continuous while EF was in place | Bridging trabeculae were present w/o radiolucent lines |

The Cochrane Collaborative risk of bias assessment demonstrated excellent inter-rater agreement (κ = 0.82). Only assessment of blinding of participants and selective reporting demonstrated discrepancy (κ = 0.54). The rest of the sources of bias, random sequence generation, allocation concealment, incomplete outcome data, and other sources of bias, were in perfect agreement (κ = 1.00) (**Table 2**). The agreed upon ‘high’ risk of bias found was in Martinez-Rondanelli *et al*.^14^ The study exhibited risk of selective reporting in that it did not report patient compliance with the removable PEMF device, which could affect the result of the study. The reviewers agreed upon an “unclear” risk of bias for blinding of participants in Itoh *et al*.^17^ They do not clearly state whether the study was blinded to the patients, but they do say that the physicians who made the determination of radiographic union were not part of the study. Knowing which patients were treated with ES could affect their decision making for determination of radiographic union, as reading radiographs is semi-objective.

**Table 2:** Risks of bias analysis by the two reviewers (P.J.N. and K.R.). Agreement overall was excellent at k = 0.82. Itoh *et al*. was agreed upon for “unclear” risk of bias for blinding of participants and personnel because they did not clearly state their blinding protocol. They do not describe a sham device for the treatment group. Additionally, they include a comment that criteria for radiographic union must be agreed upon by two surgeons who were not involved with the study, but they do not state if the surgeons were blinded to the arm that the patient was randomized to. Martinez-Rondanelli *et al*. was agreed upon for “high” risk of bias for selective reporting because they do not stratify results by patient compliance with the PEMF device, which could significantly affect the results of the study.

| Author | Title | Journal | Year | Random sequence generation | Allocation concealment | Blinding of participants and personnel | Incomplete outcome data | Selective reporting | Other |
| --- | --- | --- | --- | --- | --- | --- | --- | --- | --- |
| Faldini et al. | Electromagnetic bone growth stimulation in patients with femoral neck fractures treated with screws: prospective randomized double-blind study | Curr Orthop Pract | 2010 | Low/Low | Low/Low | Unclear/Low | Low/Low | Unclear/High | Low/Low |
| Hanneman et al. | The clinical and radiological outcome of pulsed electromagnetic field treatment for acute scaphoid fractures | J Bone and Joint Surg | 2012 | Low/Low | Unclear/Unclear | Low/Low | Low/Low | Low/Low | Low/Low |
| Hanneman et al. | CT scan-evaluated outcome of pulsed electromagnetic fields in the treatment of acute scaphoid fractures | Bone and Joint J | 2014 | Low/Low | Unclear/Unclear | Low/Low | Low/Low | Low/Low | Low/Low |
| Martinez-Rondanelli et al. | Electromagnetic stimulation as coadjuvant in the healing of diaphyseal femoral fractures: a randomized controlled trial | Colombia Medica | 2014 | Low/Low | Low/Low | Low/Low | Low/Low | High/High | Low/Low |
| Itoh et al. | Treatment of distal radius fractures with a wrist-bridging external fixation: the value of alternating electric current stimulation | J Hand Surg | 2008 | Low/Low | Low/Low | Unclear/Unclear | Unclear/Unclear | Unclear/Unclear | Low/Low |
| k = 0.82 |  |  |  | k = 1.00 | k = 1.00 | k = 0.54 | k = 1.00 | k = 0.54 | k = 1.00 |

For the outcome measure of time to radiographic union, the effect of ES did not reach significance between the control and ES groups (Mean difference (MD)) = −19.01, 95% CI = - 53.88 to 15.86, I^2^ = 81%) (**Figure 2**). There was also an insignificant reduction in time to radiographic union in the upper (MD = −15.86, 95% CI = −78.41 to 46.64, I^2^ =88%) and lower extremity (MD = −13.14, 95% CI = −27.64 to 1.37, I^2^ = 20%) subgroup analysis (**Figure 3**). Additionally, there was no significant difference in either the operative (MD = −39.37, 95% CI = −87.06 to 8.32, I^2^ = 87%) or non-operative (MD = 11.21, 95% CI = −25.18 to 47.59, I^2^ = 20%) subgroup (**Figure 4**).

**Figure 2:**
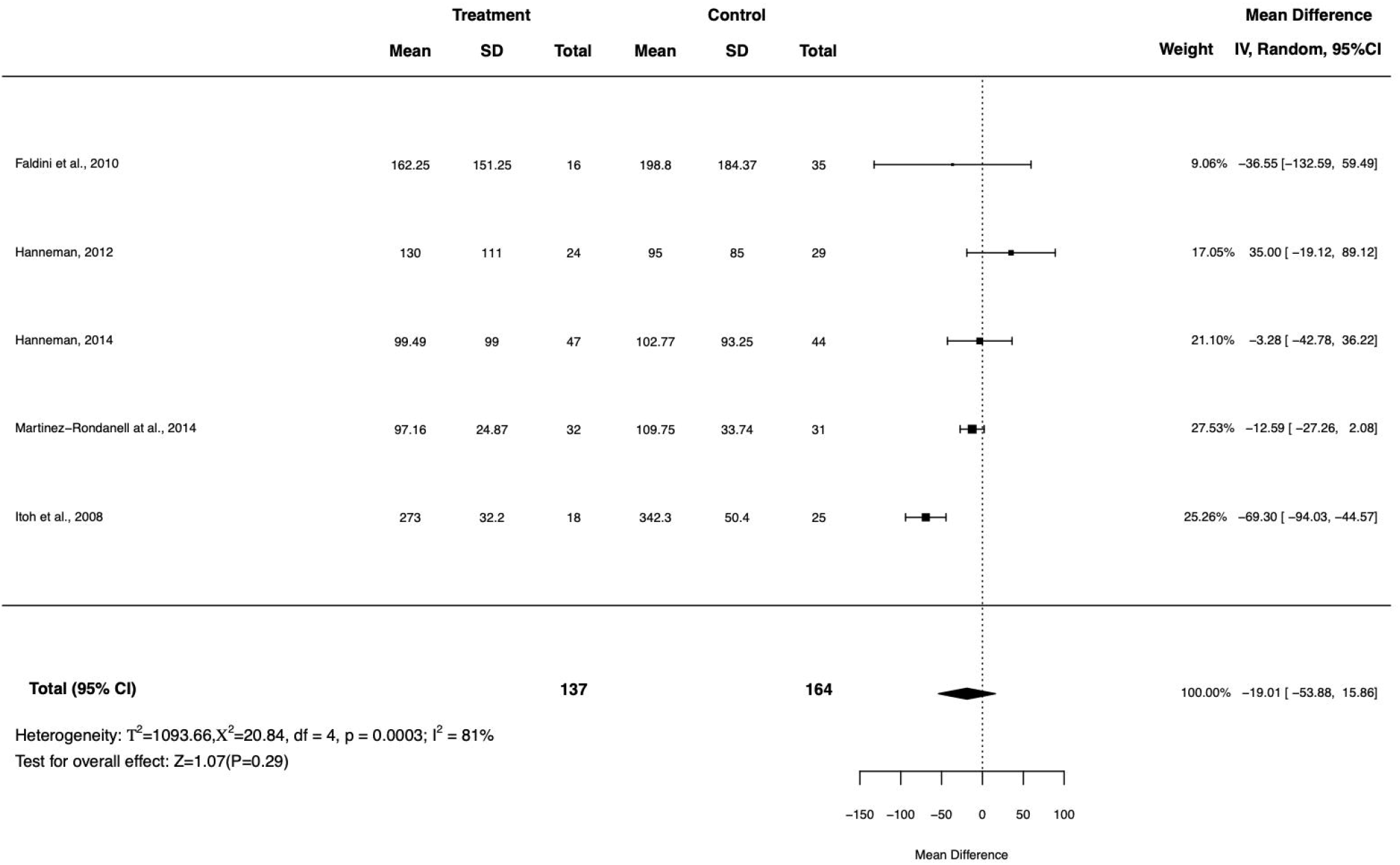
Forest plot of effect of ES on time to radiographic union in days with aggregate mean difference at the bottom of the figure. Only 1 of 5 studies demonstrated a significant difference in the treatment group compared to controls (Itoh *et al*.). Taken together, the effect of ES was determined to be not statistically different from controls.

**Figure 3:**
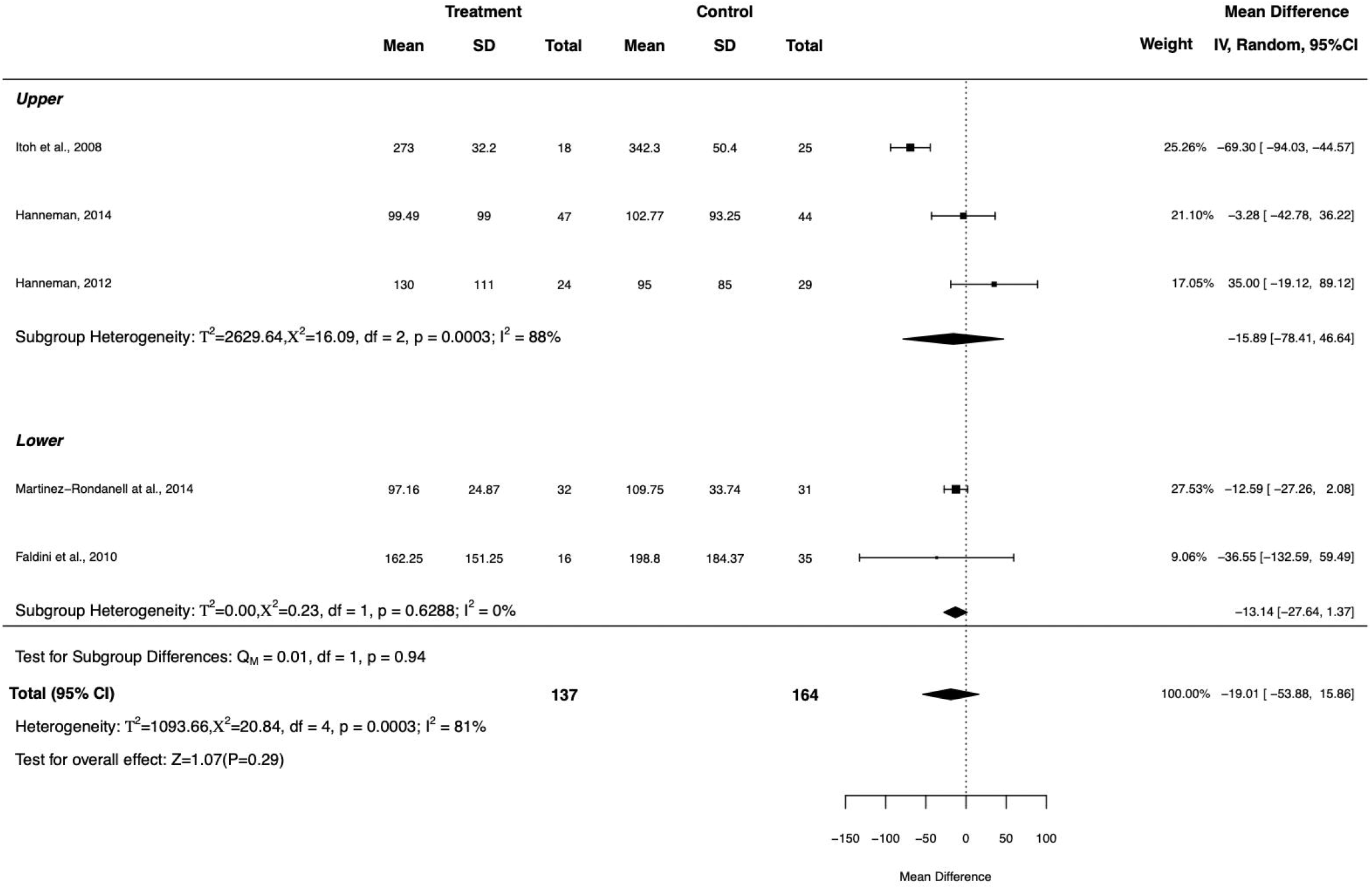
Forest plots of the upper and lower extremity subgroups of the analysis with aggregate mean difference below each Forest plot. Neither subgroup analysis reached significance.

**Figure 4:**
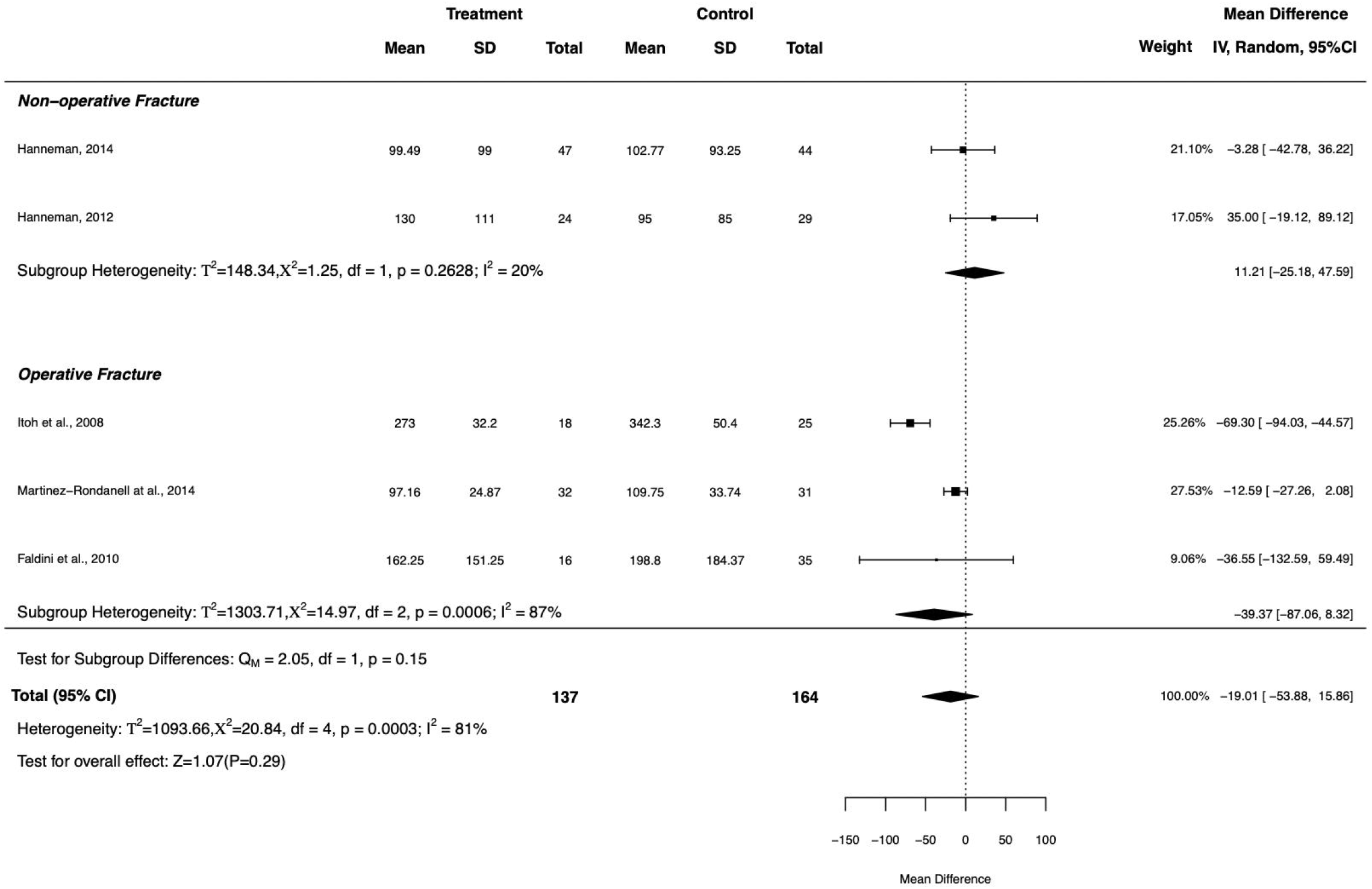
Forest plots of the operative and non-operative subgroups of the analysis with aggregate mean difference below each Foster plot. Neither subgroup analysis reached significance.

## Discussion

There were significant heterogeneities (I^2^ > 50%) reported in the overall meta-analysis (I^2^= 81%) and the upper extremity (I^2^ = 88%) and operative (I^2^ = 87%) subgroups. The high heterogeneity is likely due to the fact that some fractures healed without issue within a period of time that was considered normal for that particular fracture. However, some fractures in both the control and treatment arms of the included studies exhibited delayed healing attributable to patient factors like fracture severity, nicotine use, and corticosteroid use. This is not to say that these factors, which caused some patients in each group to heal slower than normal, affected the results of the studies because Hanneman *et al*. 2012 and 2014^15,16^ and Martinez-Rondanelli *et al*.^14^ have tables demonstrating that the rate of comorbidities known to affect fracture healing are similar between the treatment and control groups.

While Hanneman *et al*. 2012^16^ used a cast-implanted PEMF device study for treatment of scaphoid fractures and demonstrated an insignificant *increase* in time to healing in their treatment group (MD = 35.00, 95% CI = −19.12 to 89.12), the paper raised concerns about the reproducibility and inter-rater variability of radiographs for detecting union of the scaphoid.^16^ For this reason, this group repeated a similar study using computed tomography scans instead of standard radiographs for evidence of union. This repeated study demonstrated an insignificant *decrease* in time to radiographic healing (MD = −3.28, 95% CI = −42.78 to 36.22).^15^ These findings demonstrate the importance of consistent radiographic endpoints of union and sensitivity of imaging modalities.

The only study with statistically significant data for time to radiographic healing was Itoh *et al*.^17^ that used a 2Hz alternating current through a semi-invasive method of delivery (MD = - 69.30, 95% CI = −94.03 to −44.57). As stated previously, Itoh *et al*.^17^ used a spanning fixator for intra-articular distal radius fractures with a pin into the radial styloid, delivering alternating current directly into the fracture site.

Studies investigating removable PEMF devices like the Faldini *et al*.^13^ were limited by patient compliance issues with 53.3% of the treatment group reportedly using the transcutaneous PEMF device the prescribed 8 hours per day for 13 weeks. However, Hanneman *et al*.^15,16^ implanted the PEMF device within the cast for the non-operatively managed scaphoid fractures, eliminating patient compliance issues, yet there was still no significant reduction in time to healing.

In light of the robust data in *in vitro* and animal studies demonstrating augmentation of bone formation with electrical stimulation and the clinical success of the Itoh *et al*. study where an invasive method of stimulation was used,^4-6,18^ one explanation for the results of this meta-analysis could be issues with delivery of the stimulation to the fracture site. The majority of animal studies to date use an invasive means of delivering current, and the most common method of ES in human studies is PEMF.^6^ In our meta-analysis, the study utilizing invasive means of continuous current directly into the fracture site was the only study that demonstrated significant reduction in time to radiographic union (MD = −69.30, 95% CI = −94.03 to −44.57).^17^

One possible explanation for the inconsistent results in human clinical trials for ES compared to results in animals and cell cultures is a function of scale. *In vitro* and animal studies have demonstrated that proliferation of bone/osteoblast cells has a dose-dependent response to the application of a capacitively coupled electric field. Greater electrical field strengths cause a greater proliferation of cells.^19-22^ However, the fall off of an electric field from the signal source is very steep, ∼1/r^2^ where r is the distance from the electrodes.^23^

Similarly, distance to target impacts the resolution of a pulse electromagnetic field into current within tissues. The difference in distance and coverage of the electric field at the fracture site would differ greatly between humans and small animal studies based on the width of the bone and thickness of soft tissue.^24^ This may also explain why implanting an electrode did significantly decrease the time to radiographic union, which would not be subject to the same issues of scale.

Additionally, special care must be taken when calibrating the PEMF device to ensure that the fracture site is being exposed to the expected intensity of magnetic and electric field. Lunt^24^ highlights the many variables present that may alter the current generated at the fracture site. In addition to the width of the bone, the difference in resistivity of air, soft tissue, and bone causes a significant fall off in current generated in the fracture site when compared to theoretical and laboratory models.^24^ This raises another issue which may explain the negative results in the PEMF stimulator studies like the ones included. For example, another randomized, sham-controlled trial found that PEMF device had no effect on the rate of revision surgery for delayed union or non-union tibial shaft fractures.^25^ In this study, the PEMF device was strapped onto the exterior of either a plaster or fiberglass splint by the patient. There was no mention of specific calibration for each patient in the device preparation protocol. According to Lunt’s calculations,^24^ altering the geometry, thickness, and resistivity of the splint material, patients’ soft tissues, and bone would proportionally affect the current generated within the fracture site.

Therefore, the expected current within the fracture site is not as predictable as one would assume. While invasive methods of ES are not feasible in many clinical scenarios due to a significant infection risk,^26^ more research at the basic science and clinical levels is necessary to develop an effective methodology and paradigm for stimulating the fracture site *in vivo*.

One potential limitation of our study is that while Itoh et al.^17^ described more robust callus formation and improved radial height in the ES group, they used time of external fixator removal as a surrogate for time to radiographic union. The timing of external fixator removal can be affected by surgeon availability and operating room scheduling and could therefore alter the results of the study. Additionally, the study kept the external fixators in place for 39 and 48 weeks in the control and treatment arms, respectively. These are well above the normal time of external fixation for these types of fractures at our institution, which would typically be on the order of 4 to 8 weeks. And so, there is some question of the external validity of these results when applied to practice patterns within the United States. Finally, it is difficult to compare the results of the included studies as they apply ES with drastically different ES protocols and specifications. For example, the Hanneman *et al*. studies^15,16^ used PEMF stimulators that were kept on continuously while the cast was in place. In contrast, Martinez-Rondanelli *et al*.^14^ started ES by means of a PEMF stimulator 6 weeks after fixation for 1 hour per day for 8 weeks. This difference emphasizes that further studies are required to determine the optimal protocol for ES for the purpose of acute fracture healing.

No statistically significant effect was demonstrated by ES for the purposes of decreasing time to radiographic union in acute fractures. More robust RCTs with specific methodologies to address issues with patient compliance with clear and reproducible definitions of radiographic union are needed. Additionally, new developments in methods of safe and efficacious delivery of ES to fracture sites may show more robust results.

## Data Availability

The authors confirm that the data supporting the findings of this study are available within the article or its supplementary materials

## Appendix 1

### Search Strategy

((“electric stimulation”[MeSH Terms] OR (“electric”[All Fields] AND “stimulation”[All Fields]) OR “electric stimulation”[All Fields]) AND ((“bone and bones”[MeSH Terms] OR (“bone”[All Fields] AND “bones”[All Fields]) OR “bone and bones”[All Fields] OR “bone”[All Fields]) AND (“healed”[All Fields] OR “wound healing”[MeSH Terms] OR (“wound”[All Fields] AND “healing”[All Fields]) OR “wound healing”[All Fields] OR “healing”[All Fields] OR “healings”[All Fields] OR “heals”[All Fields]))) OR ((“electric stimulation”[MeSH Terms] OR (“electric”[All Fields] AND “stimulation”[All Fields]) OR “electric stimulation”[All Fields]) AND (“fractur”[All Fields] OR “fractural”[All Fields] OR “fracture s”[All Fields] OR “fractures, bone”[MeSH Terms] OR (“fractures”[All Fields] AND “bone”[All Fields]) OR “bone fractures”[All Fields] OR “fracture”[All Fields] OR “fractured”[All Fields] OR “fractures”[All Fields] OR “fracturing”[All Fields])) OR ((“electric stimulation”[MeSH Terms] OR (“electric”[All Fields] AND “stimulation”[All Fields]) OR “electric stimulation”[All Fields]) AND (“fracture healing”[MeSH Terms] OR (“fracture”[All Fields] AND “healing”[All Fields]) OR “fracture healing”[All Fields])) OR ((“direct”[All Fields] OR “directed”[All Fields] OR “directing”[All Fields] OR “direction”[All Fields] OR “directional”[All Fields] OR “directions”[All Fields] OR “directivities”[All Fields] OR “directivity”[All Fields] OR “directs”[All Fields]) AND (“current”[All Fields] OR “current s”[All Fields] OR “currently”[All Fields] OR “currents”[All Fields]) AND ((“bone and bones”[MeSH Terms] OR (“bone”[All Fields] AND “bones”[All Fields]) OR “bone and bones”[All Fields] OR “bone”[All Fields]) AND (“healed”[All Fields] OR “wound healing”[MeSH Terms] OR (“wound”[All Fields] AND “healing”[All Fields]) OR “wound healing”[All Fields] OR “healing”[All Fields] OR “healings”[All Fields] OR “heals”[All Fields]))) OR ((“inductive”[All Fields] OR “inductively”[All Fields]) AND (“couple s”[All Fields] OR “coupled”[All Fields] OR “coupling”[All Fields] OR “couplings”[All Fields] OR “family characteristics”[MeSH Terms] OR (“family”[All Fields] AND “characteristics”[All Fields]) OR “family characteristics”[All Fields] OR “couple”[All Fields] OR “couples”[All Fields]) AND ((“bone and bones”[MeSH Terms] OR (“bone”[All Fields] AND “bones”[All Fields]) OR “bone and bones”[All Fields] OR “bone”[All Fields]) AND (“healed”[All Fields] OR “wound healing”[MeSH Terms] OR (“wound”[All Fields] AND “healing”[All Fields]) OR “wound healing”[All Fields] OR “healing”[All Fields] OR “healings”[All Fields] OR “heals”[All Fields]))) OR ((“pulse”[MeSH Terms] OR “pulse”[All Fields] OR “heart rate”[MeSH Terms] OR (“heart”[All Fields] AND “rate”[All Fields]) OR “heart rate”[All Fields] OR “pulses”[All Fields] OR “pulse s”[All Fields] OR “pulsed”[All Fields] OR “pulsing”[All Fields]) AND (“electromagnetic phenomena”[MeSH Terms] OR (“electromagnetic”[All Fields] AND “phenomena”[All Fields]) OR “electromagnetic phenomena”[All Fields] OR “electromagnetic”[All Fields] OR “electromagnetics”[All Fields] OR “electromagnetic radiation”[MeSH Terms] OR (“electromagnetic”[All Fields] AND “radiation”[All Fields]) OR “electromagnetic radiation”[All Fields] OR “electromagnetism”[All Fields] OR “electromagnetical”[All Fields] OR “electromagnetically”[All Fields] OR “magnets”[MeSH Terms] OR “magnets”[All Fields] OR “electromagnet”[All Fields] OR “electromagnets”[All Fields]) AND (“epidemiology”[MeSH Subheading] OR “epidemiology”[All Fields] OR “frequency”[All Fields] OR “epidemiology”[MeSH Terms] OR “frequence”[All Fields] OR “frequences”[All Fields] OR “frequencies”[All Fields]) AND ((“bone and bones”[MeSH Terms] OR (“bone”[All Fields] AND “bones”[All Fields]) OR “bone and bones”[All Fields] OR “bone”[All Fields]) AND (“healed”[All Fields] OR “wound healing”[MeSH Terms] OR (“wound”[All Fields] AND “healing”[All Fields]) OR “wound healing”[All Fields] OR “healing”[All Fields] OR “healings”[All Fields] OR “heals”[All Fields])))

## Notes

### Competing Interest Statement

The authors have declared no competing interest.

### Funding Statement

Each author certifies that he or she has no commercial associations (eg, consultancies, stock ownership, equity interest, patent/licensing arrangements, etc) that might pose a conflict of interest in connection with the submitted article.

### Author Declarations

The paper is a meta-analysis and not required to be reviewed by the IRB

## References

1. Yelin E, Weinstein S, King T. The burden of musculoskeletal diseases in the United States. Paper presented at: Seminars in arthritis and rheumatism2016.

2. Cook JJ, Summers NJ, Cook EA. Healing in the new millennium: bone stimulators: an overview of where we’ve been and where we may be heading. Clin Podiatr Med Surg. 2015;32:45–59.

3. Fukada E, Yasuda I. On the piezoelectric effect of bone. Journal of the physical society of Japan. 1957;12:1158–1162.

4. Leppik L, Zhihua H, Mobini S, et al. Combining electrical stimulation and tissue engineering to treat large bone defects in a rat model. Sci Rep. 2018;8:6307.

5. Leppik L, Bhavsar MB, Oliveira KMC, et al. Construction and Use of an Electrical Stimulation Chamber for Enhancing Osteogenic Differentiation in Mesenchymal Stem/Stromal Cells In Vitro. J Vis Exp. 2019;

6. Bhavsar MB, Han Z, DeCoster T, et al. Electrical stimulation-based bone fracture treatment, if it works so well why do not more surgeons use it? European Journal of Trauma and Emergency Surgery. 2020;46:245–264.

7. Chang K-V, Hung C-Y, Chen W-S, et al. Effectiveness of bisphosphonate analogues and functional electrical stimulation on attenuating post-injury osteoporosis in spinal cord injury patients-a systematic review and meta-analysis. PloS one. 2013;8:e81124.

8. Aleem IS, Aleem I, Evaniew N, et al. Efficacy of Electrical Stimulators for Bone Healing: A Meta-Analysis of Randomized Sham-Controlled Trials. Sci Rep. 2016;6:31724.

9. Moher D, Liberati A, Tetzlaff J, et al. Preferred reporting items for systematic reviews and meta-analyses: the PRISMA statement. PLoS med. 2009;6:e1000097.

10. Higgins JP, Thomas J, Chandler J, et al. Cochrane handbook for systematic reviews of interventions. John Wiley & Sons; 2019.

11. Higgins JP, Thompson SG. Quantifying heterogeneity in a metaLJanalysis. Statistics in medicine. 2002;21:1539–1558.

12. Viechtbauer W. Conducting meta-analyses in R with the metafor package. Journal of statistical software. 2010;36:1–48.

13. Faldini C, Cadossi M, Luciani D, et al. Electromagnetic bone growth stimulation in patients with femoral neck fractures treated with screws: prospective randomized double-blind study. Current Orthopaedic Practice. 2010;21:282–287.

14. Martinez-Rondanelli A, Martinez JP, Moncada ME, et al. Electromagnetic stimulation as coadjuvant in the healing of diaphyseal femoral fractures: a randomized controlled trial. Colombia Médica. 2014;45:67–71.

15. Hannemann P, Van Wezenbeek M, Kolkman K, et al. CT scan-evaluated outcome of pulsed electromagnetic fields in the treatment of acute scaphoid fractures: a randomised, multicentre, double-blind, placebo-controlled trial. The Bone & Joint Journal. 2014;96:1070–1076.

16. Hannemann P, Göttgens K, van Wely B, et al. The clinical and radiological outcome of pulsed electromagnetic field treatment for acute scaphoid fractures: a randomised double-blind placebo-controlled multicentre trial. The Journal of bone and joint surgery British volume. 2012;94:1403–1408.

17. Itoh S, Ohta T, Sekino Y, et al. Treatment of distal radius fractures with a wrist-bridging external fixation: the value of alternating electric current stimulation. J Hand Surg Eur Vol. 2008;33:605–608.

18. Khalifeh JM, Zohny Z, MacEwan M, et al. Electrical Stimulation and Bone Healing: A Review of Current Technology and Clinical Applications. IEEE Rev Biomed Eng. 2018;11:217–232.

19. Griffin M, Bayat A. Electrical stimulation in bone healing: critical analysis by evaluating levels of evidence. Eplasty. 2011;11:e34.

20. Brighton CT, Okereke E, Pollack SR, et al. In vitro bone-cell response to a capacitively coupled electrical field. The role of field strength, pulse pattern, and duty cycle. Clin Orthop Relat Res. 1992:255–262.

21. Korenstein R, Somjen D, Fischler H, et al. Capacitative pulsed electric stimulation of bone cells. Induction of cyclic-AMP changes and DNA synthesis. Biochim Biophys Acta. 1984;803:302–307.

22. Wang Z, Clark CC, Brighton CT. Up-regulation of bone morphogenetic proteins in cultured murine bone cells with use of specific electric fields. J Bone Joint Surg Am. 2006;88:1053–1065.

23. Plonsey R, Barr R. Electric field stimulation of excitable tissue. IEEE transactions on biomedical engineering. 1995;42:329–336.

24. Lunt M. Theoretical model for investigating the magnetic and electric fields produced during pulsed magnetic field therapy for nonunion of the tibia. Medical and Biological Engineering and Computing. 1985;23:293–300.

25. Adie S, Harris IA, Naylor JM, et al. Pulsed electromagnetic field stimulation for acute tibial shaft fractures: a multicenter, double-blind, randomized trial. JBJS. 2011;93:1569–1576.

26. Haddad JB, Obolensky AG, Shinnick P. The biologic effects and the therapeutic mechanism of action of electric and electromagnetic field stimulation on bone and cartilage: new findings and a review of earlier work. J Altern Complement Med. 2007;13:485–490.

